# Machine Learning Prediction of Progression in FEV_1_ in the COPDGene Study

**DOI:** 10.1101/2022.01.10.22268804

**Authors:** Adel Boueiz, Zhonghui Xu, Yale Chang, Aria Masoomi, Andrew Gregory, Sharon M. Lutz, Dandi Qiao, James D. Crapo, Jennifer G. Dy, Edwin K. Silverman, Peter J. Castaldi, for the COPDGene investigators

## Abstract

**Background:** The heterogeneous nature of COPD complicates the identification of the predictors of disease progression and consequently the development of effective therapies. We aimed to improve the prediction of disease progression in COPD by using machine learning and incorporating a rich dataset of phenotypic features.

**Methods:** We included 4,496 smokers with available data from their enrollment and 5-year follow-up visits in the Genetic Epidemiology of COPD (COPDGene) study. We constructed supervised random forest models to predict 5-year progression in FEV_1_ from 46 baseline demographic, clinical, physiologic, and imaging features. Using cross-validation, we randomly partitioned participants into training and testing samples. We also validated the results in the COPDGene 10-year follow-up visit.

**Results:** Predicting the change in FEV_1_ over time is more challenging than simply predicting the future absolute FEV_1_ level. Nevertheless, the area under the ROC curves for the prediction of subjects in the top quartile of observed disease progression was 0.70 in the 10-year follow-up data. The model performance accuracy was best for GOLD1-2 subjects and it was harder to achieve accurate prediction in advanced stages of the disease. Predictive variables differed in their relative importance as well as for the predictions by GOLD grade.

**Conclusion:** This state-of-the art approach along with deep phenotyping predicts FEV_1_ progression with reasonable accuracy. There is significant room for improvement in future models. This prediction model facilitates the identification of smokers at increased risk for rapid disease progression. Such findings may be useful in the selection of patient populations for targeted clinical trials.

## INTRODUCTION

Chronic obstructive pulmonary disease (COPD) is the second leading cause of disability, the third leading cause of death, and the only major chronic disease continuing to increase in mortality ^1-4^. Novel therapies that slow disease progression could result in an improvement in COPD patients’ health status and have a substantial impact on healthcare utilization. The development of such therapies will be aided by improved tools for predicting disease progression, enabling the selection of high-risk groups for targeted treatment.

Predictive models incorporate multiple sources of information to make patient-specific predictions and are widely used in multiple areas of medical practice. Existing models of disease progression in COPD have been limited in the scope of variables assessed ^5-9^. COPD exhibits significant variation in clinical and radiographic presentation as well as disease progression ^6,10-12^. This disease heterogeneity complicates the identification of the predictors of COPD progression and limits the accuracy of predictive models. Furthermore, COPD often progresses slowly over decades and true disease progression over short time periods can be difficult to detect with existing measurements.

In this study, we aimed to improve the prediction of COPD progression by applying machine learning to a rich dataset of clinical, demographic, patient-reported variables, and imaging features in the COPDGene study. We hypothesized that deep phenotyping at the initial study visit along with random forest modelling, which exploits complex non-linear relationships and interactions among the risk factors, would facilitate the prediction of the rates of disease progression as measured by FEV_1_, a key aspect of COPD.

## MATERIALS AND METHODS

### Study populations

#### Derivation cohort - COPDGene Study Visit 1 and Visit 2

We analyzed 4,496 smokers with complete CT scans and relevant covariate data at the baseline visit (Visit 1) and 5-year follow-up visit (Visit 2) in the COPDGene cohort (NCT00608764, www.copdgene.org).

#### Temporal validation cohort - COPDGene Study Visit 3

During the Phase 3 of the COPDGene Study, enrolled subjects returned for their 10-year follow-up visit. At the time of this analysis, 1,833 smokers had completed their 10-year follow-up visit and had available 10-year spirometric and radiographic data. To predict their outcome values at Year 10 (Visit 3), we entered their 5-year (Visit 2) predictor data into the models trained in the derivation cohort.

The COPDGene study design, subject enrollment, and phenotype measurements have been previously reported ^13^ and additional information is included in the Supplement.

### Outcome variables

We constructed models to predict annualized follow-up FEV_1_ and five-year changes in FEV_1_ (ΔFEV_1_). ΔFEV_1_ (mL/year) was calculated by subtracting the Visit 1 value from the Visit 2 value and dividing by the time between Visit 1 and Visit 2. Negative values represent a lower value of the outcome at Visit 2 (i.e. worsening of the disease over the 5-year period with greater loss of FEV_1_).

### Feature selection

Candidate predictors consisted of 46 baseline demographic, clinical, physiologic, and imaging variables that were available in the COPDGene population at Visit 1 and had correlation coefficients of less than 0.90 with the other variables.

### Training, testing, and validation samples

We trained a prediction model for ΔFEV_1_ in 4,496 subjects with data from COPDGene Visit 1 and Visit 2 using a nested, 10-fold cross validation (CV) procedure. The inner fold of CV was used for parameter tuning. In the outer fold, our studied derivation cohort was randomly partitioned into ten mutually exclusive subsets (folds) of approximately equal size, using nine folds for training and one fold for testing each time for ten times. This entire procedure was repeated five times to account for the random variability of the partitioning procedure and provide more accurate estimates of the performance. This repeated resampling procedure created an ensemble of fifty models over which we averaged the predictions, and we then validated the performance of this model using data from COPDGene Visit 3 that had not been used in any aspect of the model training process (temporal validation).

### Random forest supervised machine learning

Supervised random forest is an ensemble learning method that predicts outcomes by fitting a series of decision trees and aggregating the results across trees. This method can capture non-linear dependencies and has been shown to perform well for a range of tasks ^14^. It begins building each tree by randomly selecting participants for the tree with replacement (bootstrap samples). Participants not selected in bootstrapping represent the out-of-bag set. For each bootstrap sample, a decision tree is trained by recursive binary partition of the data until the minimum node size is reached. At each node split, an optimal feature (and its split-point) is identified from a randomly selected subset of features by minimizing a loss measure. The random selection of features reduces the correlation between trees, leading to variance reduction and improved generalization performance. It also allows a moderately informative feature to assert its importance to the prediction. Once an ensemble of trees are grown, the prediction for a new sample is made by aggregating predictions (e.g. averaging for regression and majority vote for classification) from individual trees. In our study, we fixed the number of trees at 500 and tuned the hyperparameters (the bootstrap sampling fraction, the minimal node size and the number of features to use at each split) by minimizing root mean squared error (RMSE) using a nested 10-fold cross-validation within the training data.

### Random forest variable importance and their effects on the prediction

We calculated variable importance scores as the aggregated increase in the mean squared errors (IncMSE) of predictions estimated with out-of-bag samples when the values of a given variable are randomly permuted ^15,16^. The larger the increase in prediction error when permuted, the higher the variable importance score (IncMSE), and the more important the variable is to the prediction. Since the “raw” permutation importance has better statistical properties, the importance values were not normalized ^17^. Therefore, they cannot be used to compare variable importance across prediction tasks, but they can be used within the same prediction task to rank variables by their contribution to the accuracy of the final model.

### Prediction performance

We assessed the accuracy of each prediction model using the RMSE and R-squared metrics, indicators of the goodness of fit of a set of predictions to the observed values. For the prediction of ΔFEV_1_, we also assessed the ability of the models to correctly identify subjects in the top quartile of disease progression (i.e., greatest decline in FEV_1_) as quantified by the AUCs (areas under the receiver operator characteristic (ROC) curves).

### Statistical analyses

We performed a complete case analysis. Descriptive characteristics were reported respectively as percentages and medians with interquartile ranges for categorical and continuous variables. Variables were analyzed using the t-test for normally distributed variables, the Wilcoxon rank sum test for non-normally distributed variables, and chi-square tests for categorical variables. To identify differences in the quality of prediction and variable importance in subjects with different levels of COPD severity, we also constructed prediction models separately in various GOLD subgroups. All tests of significance were two-tailed with a significance threshold of P-value < 0.05.

## RESULTS

### Subject characteristics

In total, 4,496 COPDGene subjects (median age: 60; 51% men; 73% NHW) had complete phenotypic data and were included in the analysis. The participant flow diagram is shown in Figure 1.

**Figure 1.**
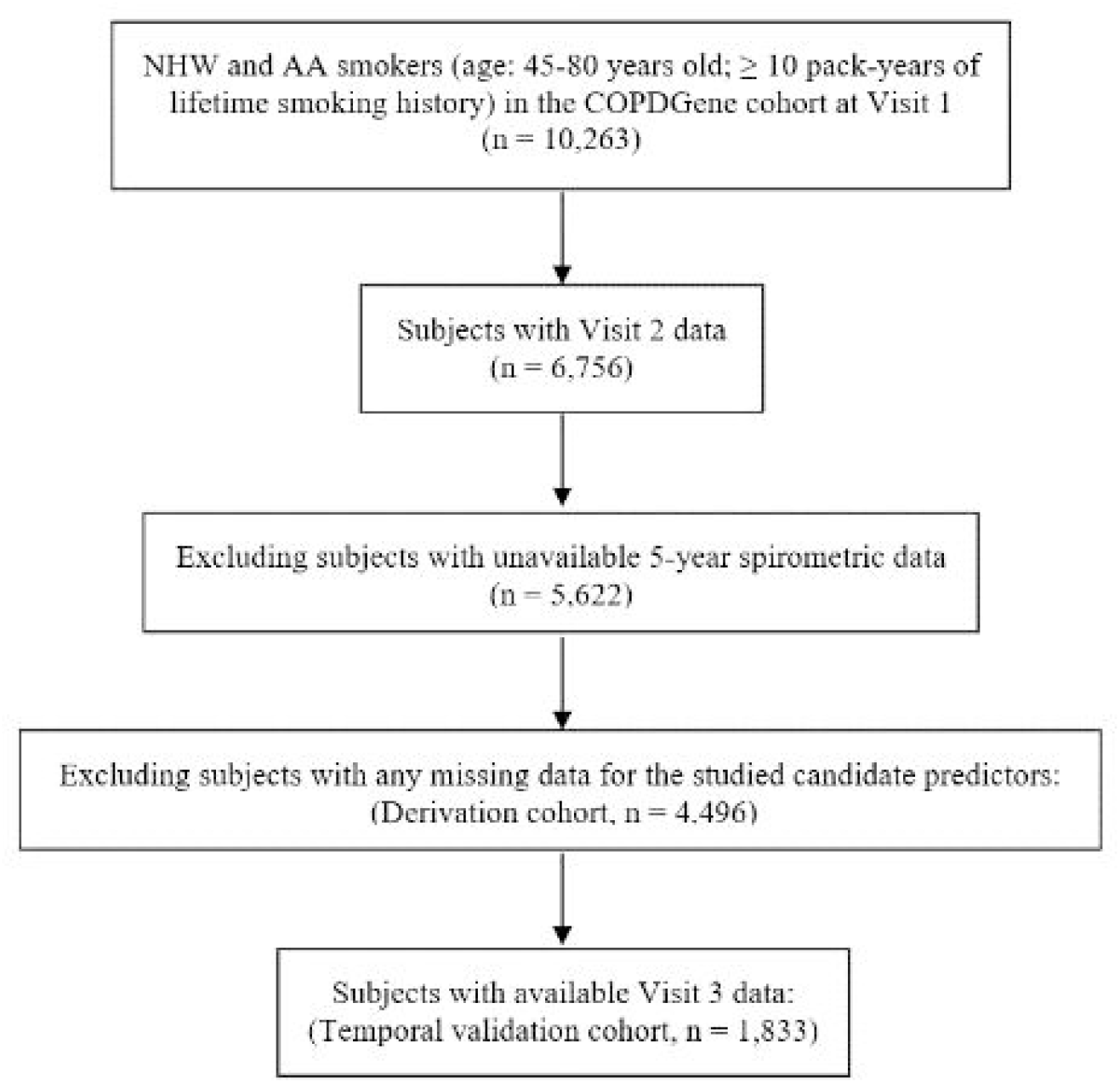
Participants’ flow diagram and general framework of the study.

### Characteristics of “rapid FEV_1_ progressors” in COPDGene

To determine the characteristics of subjects who were “rapid FEV_1_ progressors” in COPDGene, we examined the characteristics of subjects in the top quartile of progression to those in the bottom quartile (Table 1). Compared to subjects in the bottom quartile of ΔFEV_1_, those in the top quartile (“rapid FEV_1_ progressors”) had a higher proportion of males with less severe spirometric impairment at baseline but with higher exposure to smoking (pack-years and percent of current smoking), more advanced radiographic disease (total emphysema and gas trapping), more bronchodilator responsiveness, more dyspnea and chronic bronchitis symptoms, and a lower rate of obesity and metabolic syndrome.

**Table 1.**
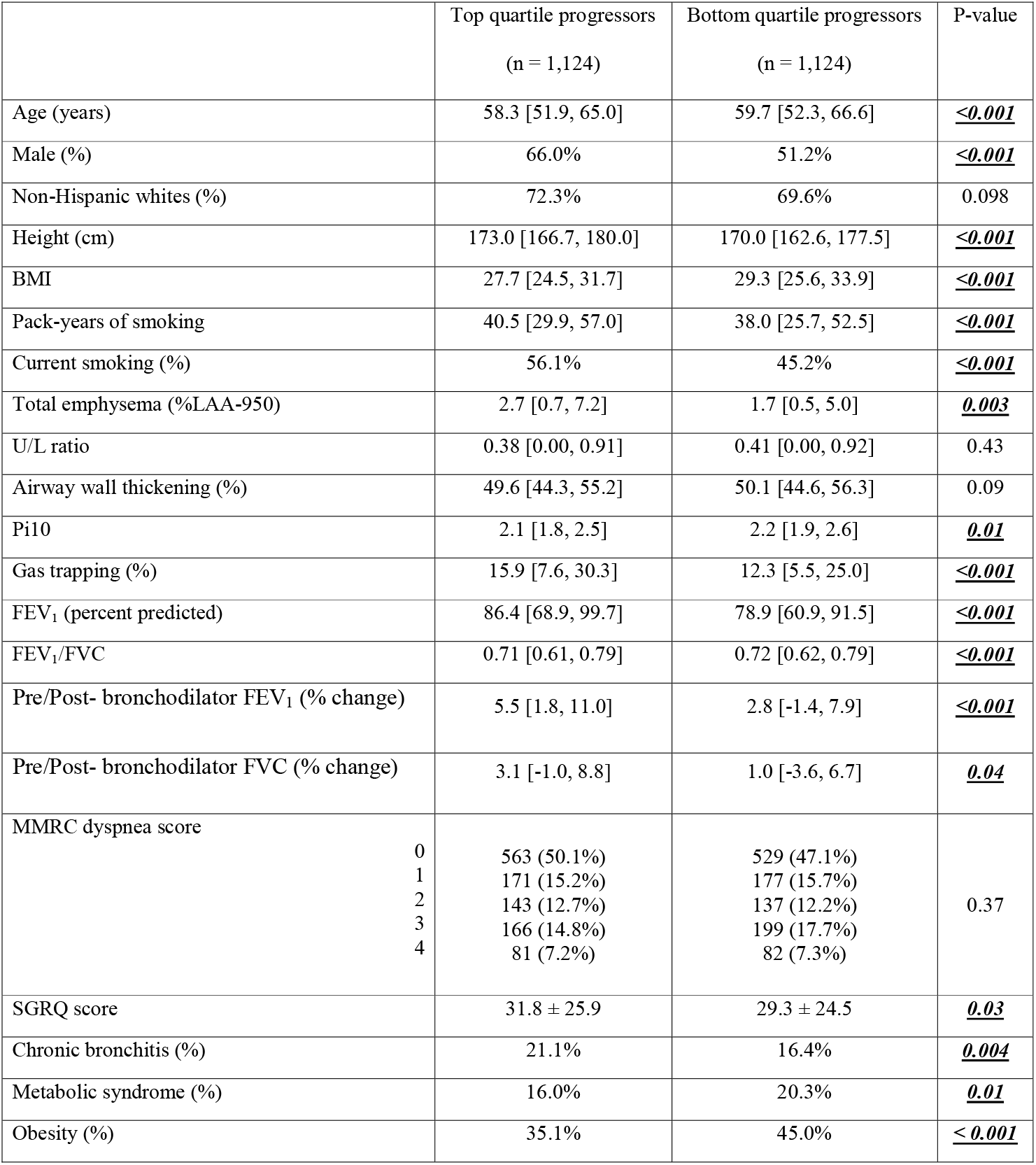
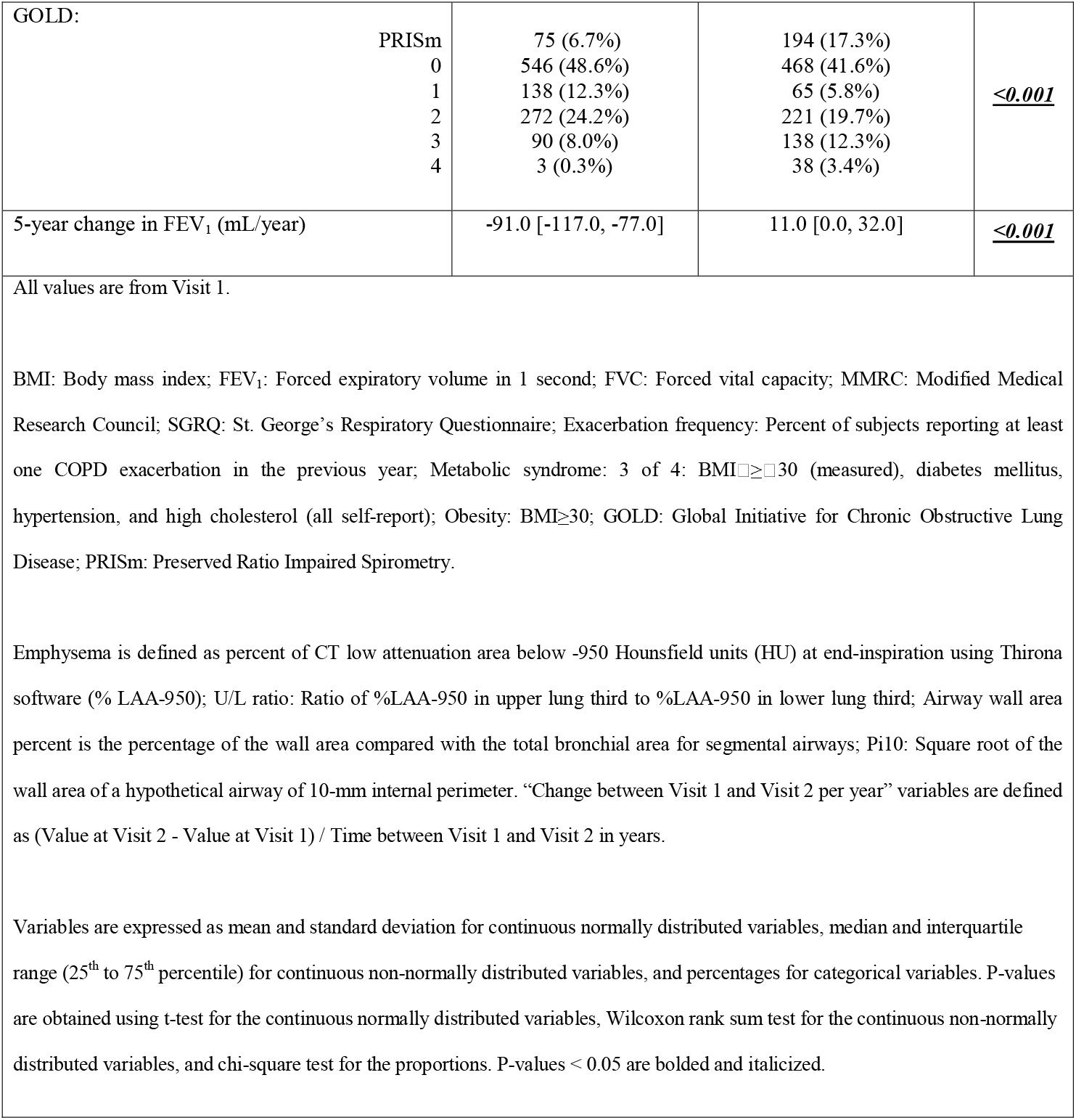
Characteristics of the rapid spirometric progressors.

The median change in FEV_1_ was -37 (IQR: -66, -9) mL/year (Figure 3). Fifty-seven percent of the studied subjects had a rate of decline in FEV_1_ of more than 30 mL/year over the 5-year period and 7% had an increase in FEV_1_ of more than 30 mL/year. Rapid FEV_1_ progressors had a median change of -91 mL/year compared to 11 mL/year for slow spirometric progressors (Table 1). When assessed according to the severity of airflow limitation, the rate of FEV_1_ decline was inversely related to the GOLD grade, with medians of ΔFEV_1_ of -46, -38, -31, -16 mL/year for GOLD 1-4, respectively.

### Prediction performance for follow-up FEV_1_ and 5-year change in FEV_1_

We constructed the prediction models using a nested cross-validation procedure and we assessed the prediction performance in the COPDGene 10-year follow-up visit. A schematic representation of our model is shown in Figure 2. The list of candidate predictors is provided in Table 2. In the cross-validation testing samples, on average, 89.6% of the variance in follow-up FEV_1_ values were explained and the area under the ROC curves for the prediction of subjects in the top quartile of observed disease progression was 0.97 (Table 3 and Figure 4). This high performance was maintained in the temporal validation with an R-squared value of 0.91 and AUC of 0.98 (Table 3). For the prediction of the change in FEV_1_ over time (ΔFEV_1_), the average R-squared value was 0.15 and AUC was 0.71 in the testing samples and respectively, 0.10 and 0.70 in the validation cohort.

**Table 2.**
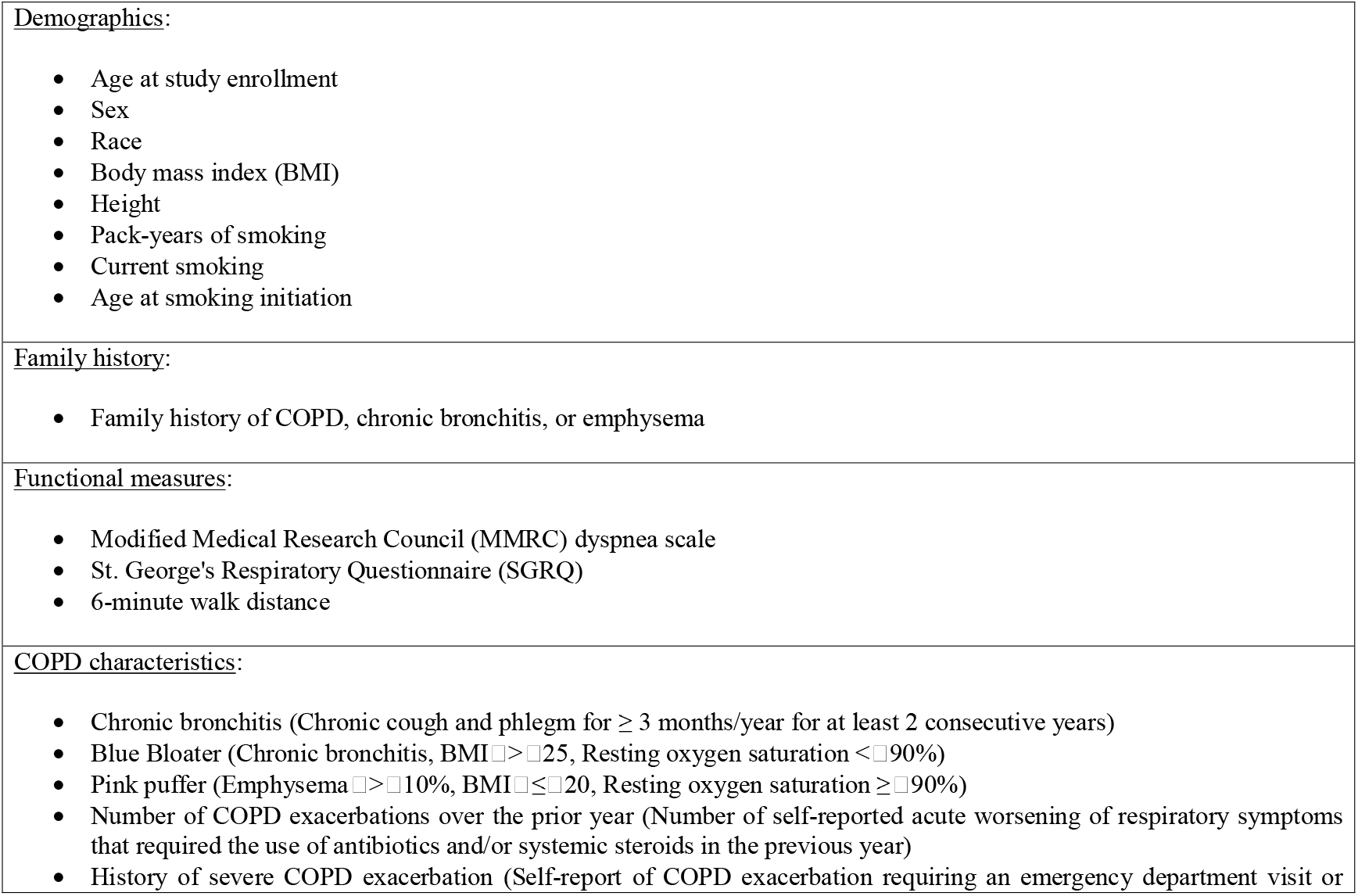
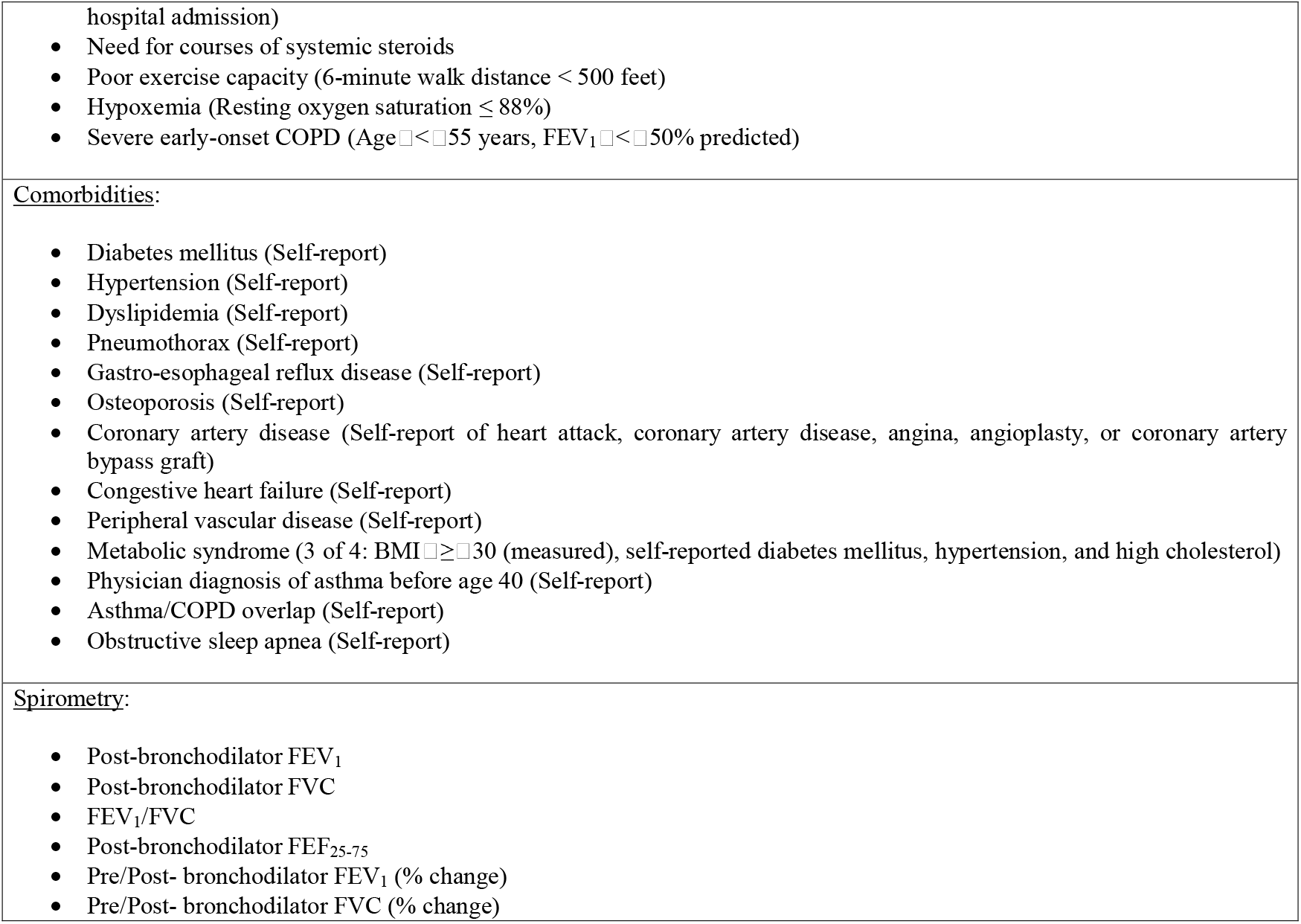
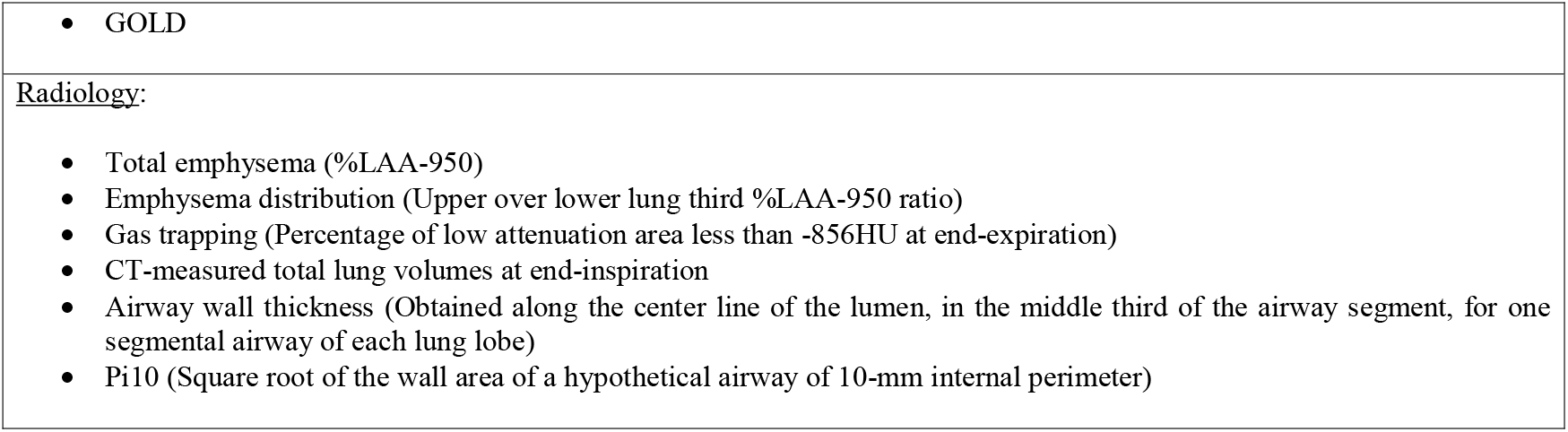
Variables included in the prediction algorithms. emographics:

**Table 3.** Prediction performance of random forest in the cross-validation testing samples and temporal validation cohort.

|  | COPDGene Visit 1 / Visit 2<br>testing samples | COPDGene Visit 2 / Visit 3<br>temporal validation cohort |
| --- | --- | --- |
| **RMSE** |  |  |
| Follow-up FEV<sub>1</sub> | 269.711 [259.252, 276.476] | 236.742 |
| Change in FEV<sub>1</sub> (mL/year) | 46.913 [45.647, 48.795] | 52.289 |
| **R-squared** |  |  |
| Follow-up FEV<sub>1</sub> | 0.896 [0.890, 0.903] | 0.913 |
| Change in FEV<sub>1</sub> (mL/year) | 0.147 [0.126, 0.173] | 0.104 |
| **AUC** |  |  |
| Follow-up FEV<sub>1</sub> | 0.974 [0.970, 0.979] | 0.975 |
| Change in FEV<sub>1</sub> (mL/year) | 0.707 [0.688, 0.724] | 0.704 |
| The derivation cohort (COPDGene Study Visit 1 and Visit 2) was randomly partitioned into training and testing samples using 10-fold cross validation. This procedure was repeated five times to account for the random variability of the partitioning procedure. This repeated resampling procedure created an ensemble of fifty models over which we averaged the predictions, and we then validated the performance of this model using data from COPDGene Visit 3 (temporal validation). To predict the outcome values at Year 10 (Visit 3), we entered the subjects' 5-year (Visit 2) predictor data into the models trained in the derivation cohort.<br>Variables are expressed as median and interquartile range (IQR) (25<sup>th</sup> to 75<sup>th</sup> percentile) when applicable.<br>AUC: Area under the ROC curve for prediction of subjects in the top quartile of COPD progression; FEV<sub>1</sub>: Forced expiratory volume in one second; RMSE: Root mean square error. |  |  |

**Figure 2.**
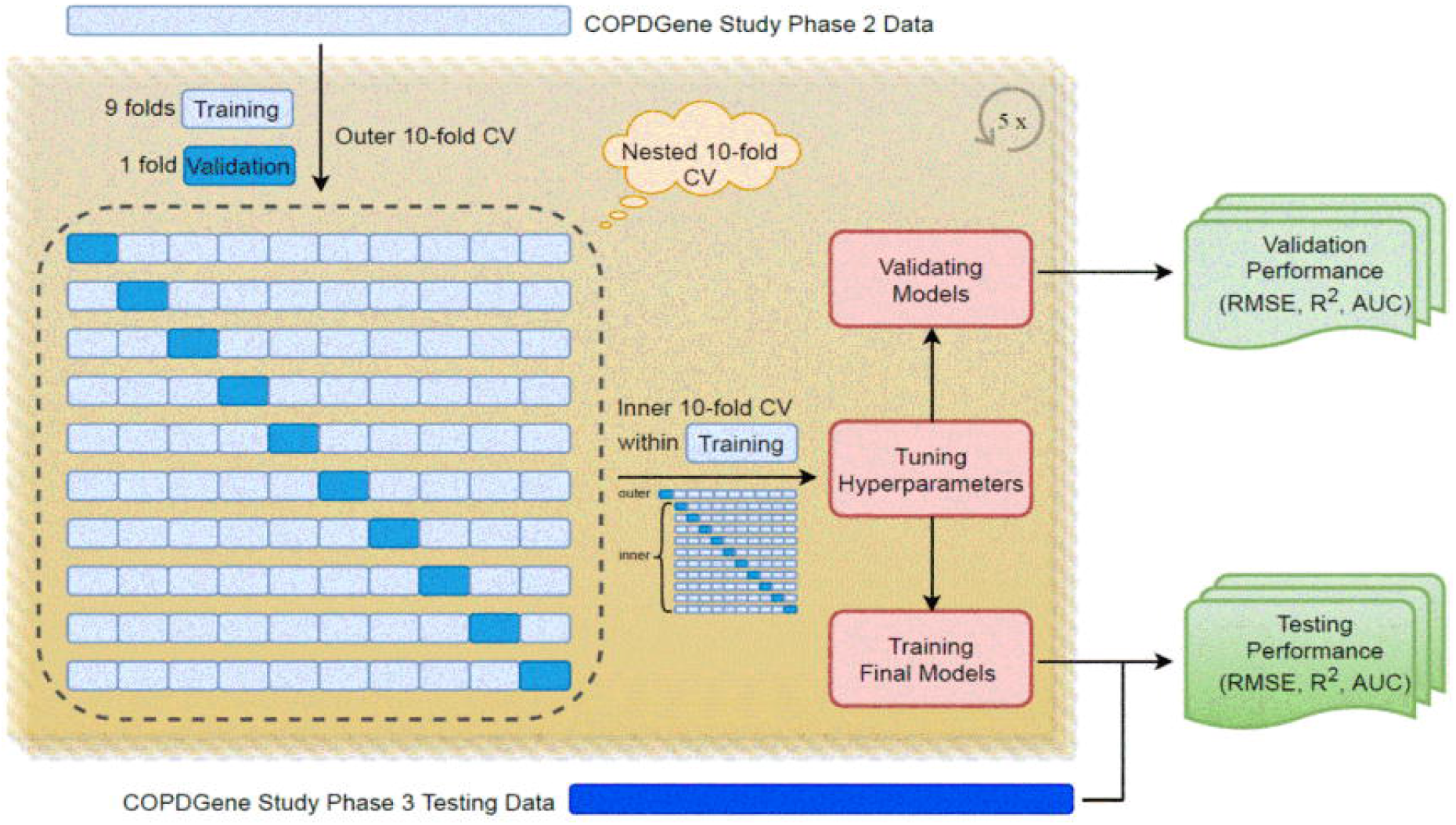
Random forest modeling framework.

**Figure 3.**
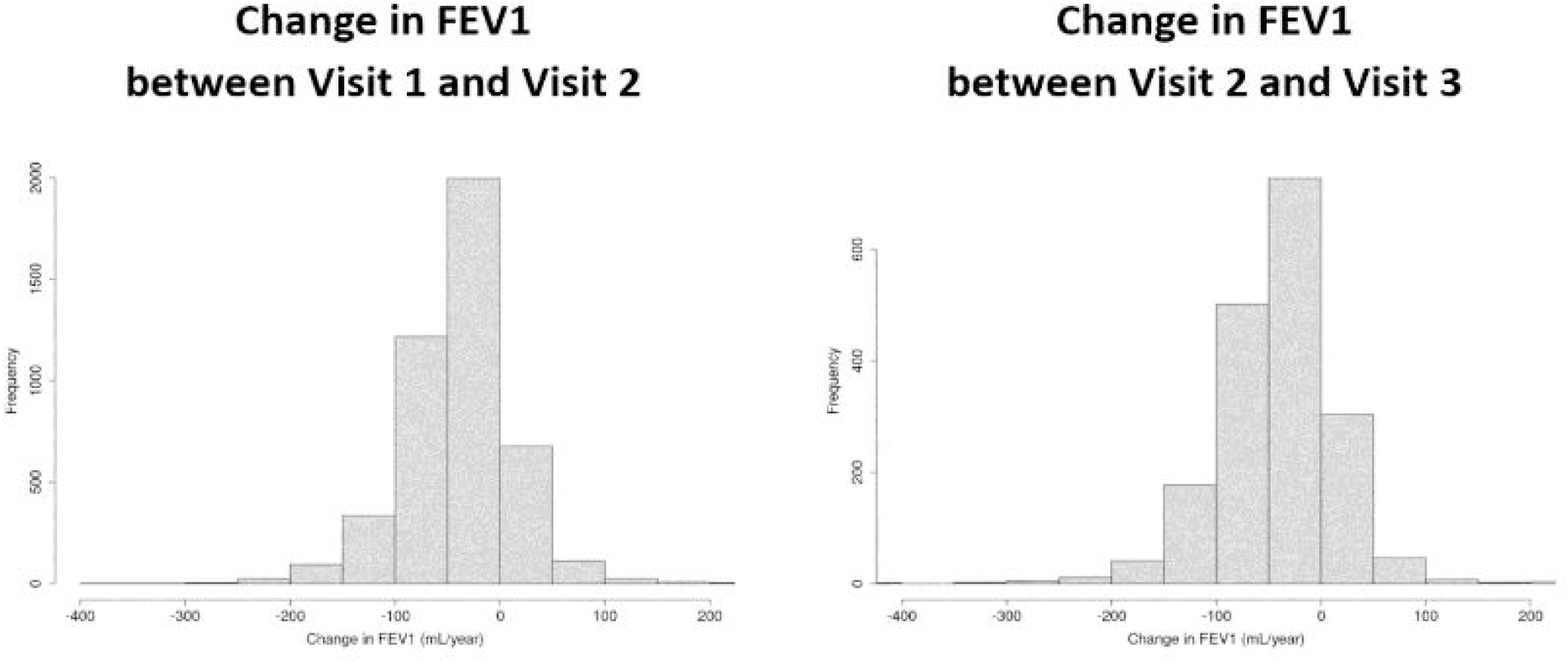
Histograms of change in FEV_1_ (ΔFEV_1_; mL/year) between Visit 1 and Visit 2 and between Visit 2 and Visit 3.

**Figure 4.**
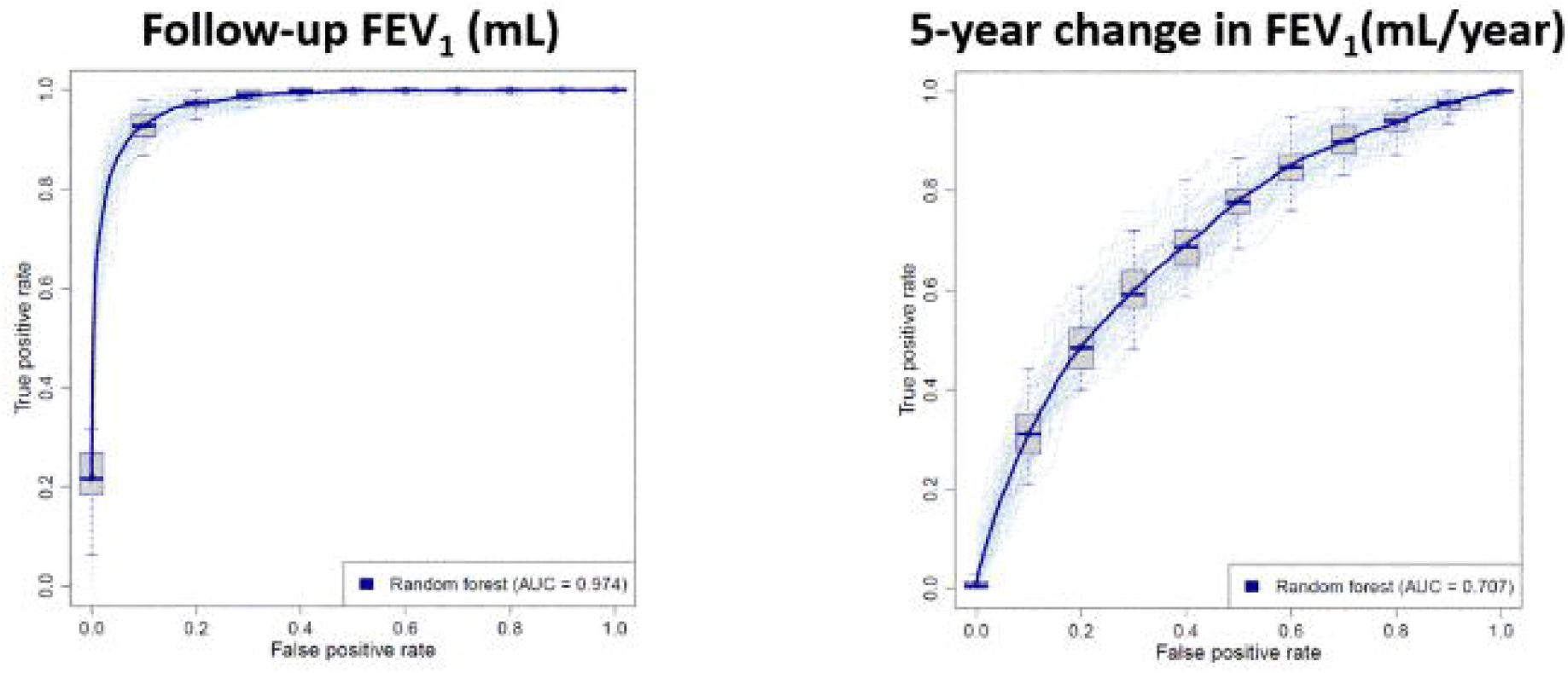
Receiver operator characteristic (ROC) curves of the performance of the random forest follow-up FEV_1_ and 5-year change in FEV_1_ models in correctly identifying subjects in the top quartiles of spirometric progression in the COPDGene Visit 1 / Visit 2 cross-validation testing samples. Solid lines represent the average performance, and colored dots represent the performance in each of the sampling iterations.

### Analysis of signal to noise ratio for 5-year change in FEV_1_

Changes in spirometric measures are more commonly used endpoints in COPD clinical trials. Predicting future FEV_1_ values is not the same as predicting the changes of FEV_1_ over the same period, since the ΔFEV_1_ over a fixed time period generally contributes a relatively small amount to the overall variance of FEV_1_ at a given time point. Given the often gradual rate of progression of COPD, five-years is a relatively short observation period, and one of the concerns is that the signal to noise ratio in our progression variables is insufficient for reliable prediction. To determine the signal-to-noise characteristics of our progression variables, we calculated the expected signal-to-noise ratio using previously published values of measurement error for FEV_1_ ^18^. An important parameter in these calculations is the extent of correlation in errors between the two study measurements. Since empiric data were unavailable, we assumed independence between these errors; therefore, these estimates likely represent a lower bound on the proportion of noise in these measures. We estimated that measurement error accounted for at least 22% of the variance of ΔFEV_1_ (calculations are included in the supplement). Thus, the theoretical upper bound for prediction performance of ΔFEV_1_ was 78%.

### Important predictors and their effects on prediction

Figure 5 shows the ranking of the top-20 predictors based on their importance scores in the random forest models. Several of the known COPD disease progression risk factors were present as top-ranked risk factors in our models and other new predictors were identified. The most important variables for FEV_1_ progression included baseline spirometry, CT-measured total lung volume, bronchodilator responsiveness, gas trapping, total emphysema, and smoking exposure. Variables like the number of COPD exacerbations in the prior year, selected comorbidities, and dyspnea scores were of less importance.

**Figure 5.**
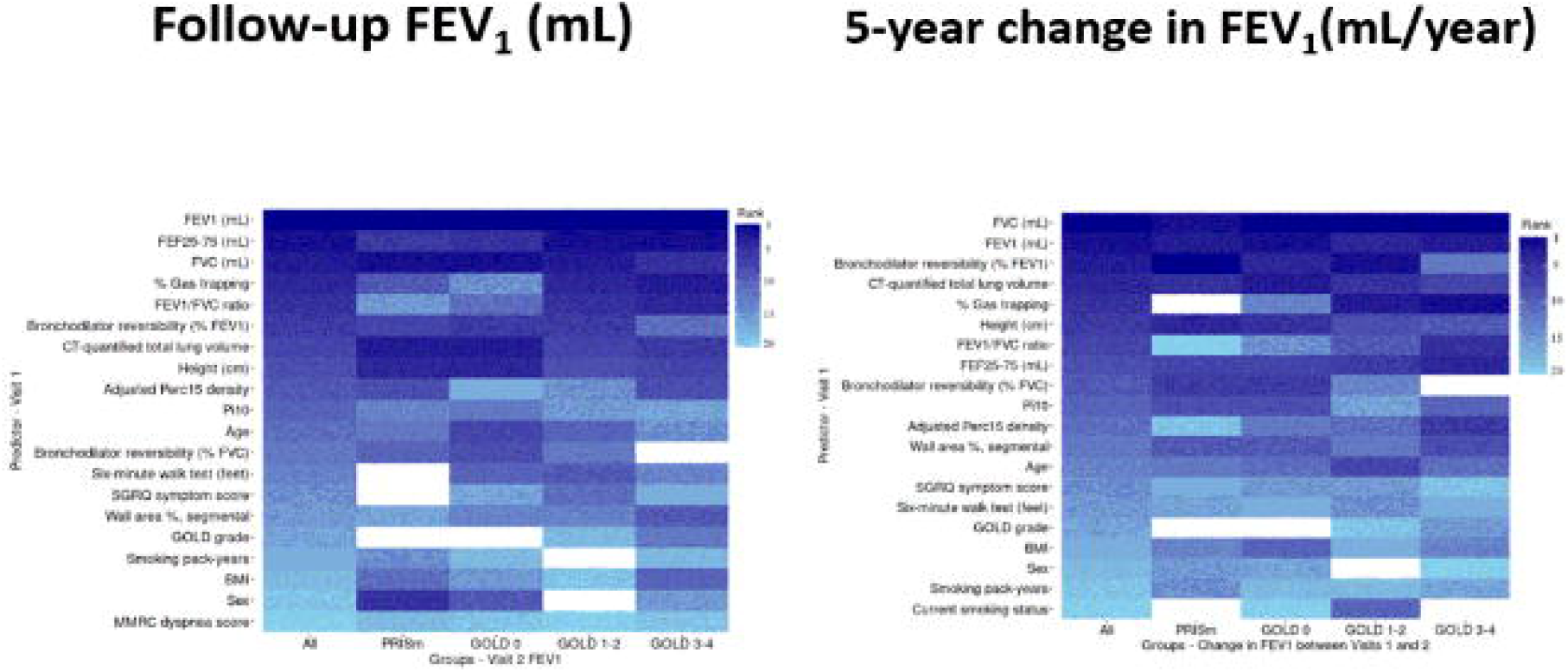
Heatmaps of the top-20 predictors of Visit 2 FEV_1_ (mL) (A) and change in FEV_1_ (mL/year) (B). The x-axis contains the group assignments (All, PRISm, GOLD0, GOLD 1-2, and GOLD 3-4). The y-axis includes the top-20 predictors ranked by their importance scores in the predictive models built in the “All” group (decreasing order with the best predictors on top). Darker shades of blue indicate a higher rank of the predictor. White cells indicate variables that do not fall within the top-20 ranks. The sample sizes were (n= 4,496 *(All)*, 499 *(PRISm)*, 2,116 *(GOLD 0)*, 1,318 *(GOLD 1-2)*, and 563 *(GOLD 3-4)*). Abbreviations: BMI: Body mass index; FEV_1_: Forced expiratory volume in 1 second; FEF25-75: Forced expiratory flow at 25–75% of forced vital capacity (FVC); Bronchodilator responsiveness (%) FEV_1_: Percentage of subjects with post-bronchodilator increase in FEV_1_ of at least 12% from baseline; Bronchodilator responsiveness (%) FVC: Percentage of subjects with post-bronchodilator increase in FVC of at least 12% from baseline; GOLD: Global Initiative for Chronic Obstructive Lung Disease; SGRQ: St. George’s Respiratory Questionnaire; MMRC: Modified Medical Research Council; Adjusted Perc15 density: Cut off value in Hounsfield units (HU) below which 15% of all voxels are distributed on a lung CT scan (per convention, adjusted Perc15 density values are reported as the HU + 1000); Gas trapping (%): Percentage of lung voxels with a density less than -856 HU at end exhalation; Pi10: Square root of the wall area of a hypothetical airway of a 10-mm internal perimeter; % Segmental airway wall thickness: Percentage of the wall relative to the total bronchial area for the segmental airways.

### Prediction of COPD progression stratified by GOLD grade

To determine whether progression was determined by different variables at different GOLD spirometric grades, we examined the performance of random forest prediction models for pre-specified subgroups of smokers stratified by GOLD grade (n= 4,496 *(Overall)*, 499 *(PRISm)*, 2,116 *(GOLD 0)*, 1,318 *(GOLD 1-2)*, and 563 *(GOLD 3-4)*). We observed significant differences in predictive performance across these subgroups. The model performance accuracy was best for GOLD 1-2 subjects and it was harder to achieve accurate prediction in advanced stages of the disease. The area under the ROC curves for the prediction of subjects in the top quartile of disease progression was 0.66 (GOLD 0), 0.73 (GOLD 1-2), and 0.58 (GOLD 3-4). The predictors of disease progression were also different by GOLD grade (Figure 5). For instance, bronchodilator responsiveness seems to be less important and emphysema and airway disease more important in the prediction of ΔFEV_1_ in subjects at more advanced stages of the disease.

## DISCUSSION

This current study showed that the prediction of change in FEV_1_, which is more relevant for disease progression, is more challenging than predicting absolute FEV_1_ level. Our prediction models for ΔFEV_1_ represent the current state of the art for prediction of prospective change in FEV_1_, but there is significant room for improvement in future models. The most important predictive variables came from a wide range of clinical, spirometric, and imaging features. Baseline spirometry, CT-measured total lung volumes, and bronchodilator responsiveness dominated the prediction. In addition, the predictive performance and the relative importance of predictors differed by GOLD grade.

Several screening tools are available to identify patients with undiagnosed COPD and to predict outcomes in patients with COPD ^1,8,9,19-24^. While Zafari et al. and Chen et al. developed and validated risk models to accurately predict lung function trajectory ^8,9^, our study is the first to apply advanced machine learning methods, use an extensive set of phenotypic measurements and comorbidities, predict not only the follow-up values but also the more relevant “change” variables, and identify the relative importance of the predictors at various stages of the disease. With respect to the outcomes evaluated in these two papers, our predictive models gave similar performance for the prediction of future values of FEV_1_. Our study added the prediction of prospective changes in FEV_1_ that were not reported in these previously published studies. Predicting the change over time is more challenging than simply predicting the future value, since the change typically represents a small proportion of the overall variance in a given pair of FEV_1_ measurements separated by five years or less. However, it is important to assess the ability of models to predict prospective changes since this is an important outcome for clinical trials.

The predictive accuracy of our models may potentially be further improved by including additional predictors (such as DLCO, pulmonary vascular measures, and relevant molecular biomarkers) and exploring other machine learning algorithms (such as deep learning). At present, these models are not ready for clinical use but could be useful in the design of COPD clinical trials to enrich the study populations by patients who are most likely to experience rapid disease progression and benefit from therapeutic interventions. For clinical use, better performing models that have been more extensively validated in multiple additional and relevant target populations are necessary.

Rapid decline in lung function has previously been associated with a range of factors such as smoking exposure, bronchodilator reversibility, higher baseline FEV_1_, higher baseline FVC, exacerbations in the prior year, low BMI, African American race, female sex, emphysema, upper lobe emphysema predominance, and CT-detected small airway abnormalities ^5,6,8,25-30^. Our study detected several of these known COPD disease progression risk factors and identified other new predictors for FEV_1_ decline. Our study is the first to our knowledge to demonstrate that the patterns of predictors vary by GOLD spirometric grade. The intriguing variations in the importance of different risk factors depending on the studied subgroup may help inform further exploration of predictive risk factors and future development of new risk prediction algorithms.

The relative unimportance of certain traditional risk factors such as COPD exacerbations in the prior year, selected comorbidities, race, and sex in our machine learning predictive models may be consistent with the disparate results from previous studies. For example, although some publications have suggested a significant excess loss of FEV_1_ for each COPD exacerbation ^26,31,32^, others have reported minimal ^6^ or no relationship ^33^. Such discrepancy may also result from differences in methodology between studies as well as differences sample size, study duration, study population, and variable definitions. The relative unimportance of certain traditional risk factors in our models may also indicate that, while these risk factors may attain statistical significance in some models, they do not provide much additional predictive value after considering more important risk factors.

This study has a number of strengths. Analyses were performed within a well-characterized cohort that included subjects at all stages of disease severity. In addition, by focusing on prediction rather than the study of individual risk factors, our results provide useful context regarding the relative importance of specific predictors. By constructing models in subjects stratified by GOLD spirometric grade, we demonstrated that patterns of optimal predictors vary by specific disease outcome and GOLD grade. Validation of our findings in the temporal cohort represents another strength of our paper.

Our study also has limitations. We only used two measurements of lung function separated by approximately 5 years. The large sample size available helped to overcome some of the inherent challenges in low signal-to-noise ratio with studies of COPD progression over a relatively short period of time. However, with longer follow-up and more measurements, we will be better able to isolate measurement noise from real disease progression which will result in greater predictive accuracy. Our analysis was based on subjects who had completed their second study visit, and it is possible that patients who were lost to follow-up differed from those available for analysis. Many of the patients with airflow obstruction were receiving therapy for their disease. Although no existing pharmacotherapy has been conclusively shown to affect the rates of disease progression, this still may have influenced our results. However, we chose not to include pharmacotherapy data in these analyses in order to reduce biases likely present in patient-reported pharmaco-epidemiologic data ^34,35^. Lastly, it is recognized that as the number of potential risk factors increases, the complexity of the models can cause overfitting. We addressed this by appropriate hyperparameter tuning and by evaluating the performance of our predictive models in cross-validation and in the temporal cohort.

Random forest machine learning in conjunction with deep phenotyping improves the prediction accuracy of COPD progression. The present study improves our ability to identify patients at risk for rapid disease progression, and these models may be useful for the development of targeted disease-modifying therapies.

## Data Availability

All data produced in the present study are available upon reasonable request to the authors.

## ABBREVIATION LIST

AUC: Area under the curve
BMI: Body mass index
COPD: Chronic Obstructive Pulmonary Disease
COPDGene study: Genetic Epidemiology of COPD study
ΔFEV_1_: Annualized five-year changes in FEV_1_
FEV_1_: Forced expiratory volume in one second
FVC: Forced vital capacity
GOLD: Global Initiative for Chronic Obstructive Lung Disease spirometric grading system
HU: Hounsfield units
IQR: Interquartile range
%LAA-950: Percent of CT scan low attenuation area below -950 HU at end-inspiration
MMRC: Modified Medical Research Council
NHW: Non-Hispanic White
RF: Random forest
RMSE: Root mean squared error
ROC: Receiver operator characteristic
SGRQ: St. George’s Respiratory Questionnaire

## Author Contributions

Drs. Boueiz and Castaldi had full access to all the data in the study and take responsibility for the integrity of the data and the accuracy of the data analysis. COPDGene investigators were instrumental in the design and implementation of the COPDGene study and collected and analyzed data cited in this article. All authors have reviewed, approved and endorsed all content and conclusions of this article.

## Declaration of Interest

The COPDGene study is funded by National Heart, Lung, and Blood Institute grants U01 HL089897 and U01 HL089856. The COPDGene study (NCT00608764) is also supported by the COPD Foundation through contributions made to an Industry Advisory Board that has included AstraZeneca, Bayer Pharmaceuticals, Boehringer-Ingelheim, Genentech, GlaxoSmithKline, Novartis, Pfizer, and Sunovion. While some individual authors of this manuscript were employed by one of the listed funders at the time the work of this study was conducted, these employment relationships did not constitute undue influence by funders. These funders have had no official role in the collection, management, analysis and interpretation of the data or design and conduct of the study. All authors have completed a Conflict of Interest form, disclosing any real or apparent financial relationships including receiving royalties, honoraria or fees for consulting, lectures, speakers’ bureaus, continuing education, medical advisory boards or expert testimony; receipt of grants; travel reimbursement; direct employment compensation. These disclosure forms have been filed with the Chronic Obstructive Pulmonary Diseases: Journal of the COPD Foundation Editorial Office and are available for review, upon request at.

## ACKNOWLEDGEMENTS

The authors wish to thank the thousands of patients who participated in the COPDGene study over the last 10 years.

**COPDGene Investigators - Core Units:**

*Administrative Center*: James D. Crapo, MD (PI); Edwin K. Silverman, MD, PhD (PI); Barry J. Make, MD; Elizabeth A. Regan, MD, PhD

*Genetic Analysis Center*: Terri H. Beaty, PhD; Peter J. Castaldi, MD, MSc; Michael H. Cho, MD, MPH; Dawn L. DeMeo, MD, MPH; Adel Boueiz, MD, MMSc; Marilyn G. Foreman, MD, MS; Auyon Ghosh, MD; Lystra P. Hayden, MD, MMSc; Craig P. Hersh, MD, MPH; Jacqueline Hetmanski, MS; Brian D. Hobbs, MD, MMSc; John E. Hokanson, MPH, PhD; Wonji Kim, PhD; Nan Laird, PhD; Christoph Lange, PhD; Sharon M. Lutz, PhD; Merry-Lynn McDonald, PhD; Dmitry Prokopenko, PhD; Matthew Moll, MD, MPH; Jarrett Morrow, PhD; Dandi Qiao, PhD; Elizabeth A. Regan, MD, PhD; Aabida Saferali, PhD; Phuwanat Sakornsakolpat, MD; Edwin K. Silverman, MD, PhD; Emily S. Wan, MD; Jeong Yun, MD, MPH

*Imaging Center*: Juan Pablo Centeno; Jean-Paul Charbonnier, PhD; Harvey O. Coxson, PhD; Craig J. Galban, PhD; MeiLan K. Han, MD, MS; Eric A. Hoffman, Stephen Humphries, PhD; Francine L. Jacobson, MD, MPH; Philip F. Judy, PhD; Ella A. Kazerooni, MD; Alex Kluiber; David A. Lynch, MB; Pietro Nardelli, PhD; John D. Newell, Jr., MD; Aleena Notary; Andrea Oh, MD; Elizabeth A. Regan, MD, PhD; James C. Ross, PhD; Raul San Jose Estepar, PhD; Joyce Schroeder, MD; Jered Sieren; Berend C. Stoel, PhD; Juerg Tschirren, PhD; Edwin Van Beek, MD, PhD; Bram van Ginneken, PhD; Eva van Rikxoort, PhD; Gonzalo Vegas SanchezFerrero, PhD; Lucas Veitel; George R. Washko, MD; Carla G. Wilson, MS

*PFT QA Center, Salt Lake City, UT*: Robert Jensen, PhD

*Data Coordinating Center and Biostatistics, National Jewish Health, Denver, CO*: Douglas Everett, PhD; Jim Crooks, PhD; Katherine Pratte, PhD; Matt Strand, PhD; Carla G. Wilson, MS

*Epidemiology Core, University of Colorado Anschutz Medical Campus, Aurora*, CO:John E. Hokanson, MPH, PhD; Erin Austin, PhD; Gregory Kinney, MPH, PhD; Sharon M. Lutz, PhD; Kendra A. Young, PhDVersion Date: March 26, 2021

*Mortality Adjudication Core*: Surya P. Bhatt, MD; Jessica Bon, MD; Alejandro A. Diaz, MD, MPH; MeiLan K. Han, MD, MS; Barry Make, MD; Susan Murray, ScD; Elizabeth Regan, MD; Xavier Soler, MD; Carla G. Wilson, MS

*Biomarker Core*: Russell P. Bowler, MD, PhD; Katerina Kechris, PhD; Farnoush BanaeiKashani, PhD

**COPDGene Investigators - Clinical Centers:**

*Ann Arbor VA*: Jeffrey L. Curtis, MD; Perry G. Pernicano, MD

*Baylor College of Medicine, Houston, TX*: Nicola Hanania, MD, MS; Mustafa Atik, MD; Aladin Boriek, PhD; Kalpatha Guntupalli, MD; Elizabeth Guy, MD; Amit Parulekar, MD

*Brigham and Women’s Hospital, Boston, MA*: Dawn L. DeMeo, MD, MPH; Craig Hersh, MD, MPH; Francine L. Jacobson, MD, MPH; George Washko, MD

*Columbia University, New York, NY*: R. Graham Barr, MD, DrPH; John Austin, MD; Belinda D’Souza, MD; Byron Thomashow, MD

*Duke University Medical Center, Durham, NC*: Neil MacIntyre, Jr., MD; H. Page McAdams, MD; Lacey Washington, MD

*HealthPartners Research Institute*, Minneapolis, MN: Charlene McEvoy, MD, MPH; Joseph Tashjian, MD

*Johns Hopkins University*, Baltimore, MD: Robert Wise, MD; Robert Brown, MD; Nadia N. Hansel, MD, MPH; Karen Horton, MD; Allison Lambert, MD, MHS; Nirupama Putcha, MD, MHS

*Lundquist Institute for Biomedical Innovation at Harbor UCLA Medical Center*, Torrance, CA: Richard Casaburi, PhD, MD; Alessandra Adami, PhD; Matthew Budoff, MD; Hans Fischer, MD; Janos Porszasz, MD, PhD; Harry Rossiter, PhD; William Stringer, MD

*Michael E. DeBakey VAMC*, Houston, TX: Amir Sharafkhaneh, MD, PhD; Charlie Lan, DO

*Minneapolis VA*: Christine Wendt, MD; Brian Bell, MD; Ken M. Kunisaki, MD, MS

*Morehouse School of Medicine, Atlanta, GA*: Eric L. Flenaugh, MD; Hirut Gebrekristos, PhD; Mario Ponce, MD; Silanath Terpenning, MD; Gloria Westney, MD, MS

*National Jewish Health, Denver, CO*: Russell Bowler, MD, PhD; David A. Lynch, MB Reliant Medical Group, Worcester, MA: Richard Rosiello, MD; David Pace, MD

*Temple University, Philadelphia, PA*: Gerard Criner, MD; David Ciccolella, MD; Francis Cordova, MD; Chandra Dass, MD; Gilbert D’Alonzo, DO; Parag Desai, MD; Michael Jacobs, PharmD; Steven Kelsen, MD, PhD; Victor Kim, MD; A. James Mamary, MD; Nathaniel Marchetti, DO; Aditi Satti, MD; Kartik Shenoy, MD; Robert M. Steiner, MD; Alex Swift, MD; Irene Swift, MD; Maria Elena Vega-Sanchez, MD

*University of Alabama, Birmingham, AL*: Mark Dransfield, MD; William Bailey, MD; Surya P. Bhatt, MD; Anand Iyer, MD; Hrudaya Nath, MD; J. Michael Wells, MD

*University of California, San Diego, CA*: Douglas Conrad, MD; Xavier Soler, MD, PhD; Andrew Yen, MD

*University of Iowa, Iowa City, IA*: Alejandro P. Comellas, MD; Karin F. Hoth, PhD; John Newell, Jr., MD; Brad Thompson, MD

*University of Michigan, Ann Arbor, MI*: MeiLan K. Han, MD MS; Ella Kazerooni, MD MS; Wassim Labaki, MD MS; Craig Galban, PhD; Dharshan Vummidi, MD

*University of Minnesota, Minneapolis, MN*: Joanne Billings, MD; Abbie Begnaud, MD; Tadashi Allen, MD

*University of Pittsburgh, Pittsburgh, PA*: Frank Sciurba, MD; Jessica Bon, MD; Divay Chandra, MD, MSc; Joel Weissfeld, MD, MPH

*University of Texas Health, San Antonio, San Antonio, TX*: Antonio Anzueto, MD; Sandra Adams, MD; Diego Maselli-Caceres, MD; Mario E. Ruiz, MD; Harjinder Singh

## Notes

**Funding Sources:** This work was supported by NHLBI K08 HL141601, R01 HL124233, R01 HL126596, R01 HL147326, U01 HL089897, and U01 HL089856. The COPDGene study (NCT00608764) is also supported by the COPD Foundation through contributions made to an Industry Advisory Board that has included AstraZeneca, Bayer Pharmaceuticals, Boehringer-Ingelheim, Genentech, GlaxoSmithKline, Novartis, Pfizer, and Sunovion.

### Competing Interest Statement

The authors have declared no competing interest.

### Funding Statement

This work was supported by NHLBI K08 HL141601, R01 HL124233, R01 HL126596, R01 HL147326, U01 HL089897, and U01 HL089856. The COPDGene study (NCT00608764) is also supported by the COPD Foundation through contributions made to an Industry Advisory Board that has included AstraZeneca, Bayer Pharmaceuticals, Boehringer-Ingelheim, Genentech, GlaxoSmithKline, Novartis, Pfizer, and Sunovion.

### Author Declarations

Institutional review board (IRB) approval was obtained. IRB Protocol Title: Genetic Epidemiology of COPD. IRB Protocol Number: Brigham and Women's Hospital / 2007P000554

