## Supplement for "Machine Learning Prediction of Progression in FEV_1_ in the COPDGene Study"

Adel Boueiz^1,2^, Zhonghui Xu^1^, Yale [Chang](http://www.ncbi.nlm.nih.gov/pubmed/?term=Chang%20Y%5BAuthor%5D&cauthor=true&cauthor_uid=26773458)^3^, Aria Masoomi^3^, Sharon M. Lutz^4^, Dandi Qiao^1^, James D. Crapo^5^, Jennifer G. Dy^3^, Edwin K. Silverman^1,2^, Peter J. Castaldi^1,6^, for the COPDGene investigators.

^1^Channing Division of Network Medicine, Brigham and Women’s Hospital, Harvard Medical School, Boston, MA; ^2^Pulmonary and Critical Care Division, Department of Medicine, Brigham and Women’s Hospital, Harvard Medical School, Boston, MA; ^3^Department of Electrical and Computer Engineering, Northeastern University, Boston, MA; ^4^Department of Population Medicine, Harvard Pilgrim Health Care Institute, Boston, MA; ^5^Division of Pulmonary Medicine, Department of Medicine, National Jewish Health, Denver, CO; ^6^Division of General Medicine and Primary Care, Brigham and Women’s Hospital, Harvard Medical School, Boston, MA.

**Supplemental methods**

***Study populations***

**COPDGene Study** (NCT00608764, www.copdgene.org). COPDGene is an ongoing multicenter study designed to investigate the genetic and epidemiologic associations of COPD ^1^. COPDGene enrolled self-identified non-Hispanic whites (NHW) and African-Americans (AA) smokers across the full spectrum of disease severity as defined by the Global Initiative for Chronic Obstructive Lung Disease (GOLD) spirometric staging system ^2^. Subjects were aged 45 to 80 years at study enrollment and had at least 10 pack-years of lifetime smoking history. They were recruited at 21 U.S. clinical centers. Exclusion criteria included pregnancy, history of other lung diseases except asthma, prior lobectomy or lung volume reduction surgery (LVRS), active cancer, or known or suspected lung cancer. Subjects who underwent LVRS or lung transplant between visits and subjects who had more than 1-liter increase of FEV_1_ between visits were also excluded from the analysis. Written, informed consents were obtained for all participants. The study and consent forms were approved by the Partners Human Research Committee (number 2007P000554/BWH).

***Demographic and clinical data***

Data on demographics, smoking burden, respiratory morbidity, exacerbations, and comorbidities used in this analysis were recorded at the baseline visit (Visit 1). History of COPD exacerbations in the previous year was defined as acute worsening of respiratory symptoms that required the use of antibiotics and/or systemic steroids ^3^. Severe exacerbation was defined as a COPD exacerbation requiring an emergency department visit or hospital admission. Respiratory disease-related health impairment and quality of life were assessed using the St George’s Respiratory Questionnaire (SGRQ) ^4^, and dyspnea was evaluated using the Modified Medical Research Council (MMRC) dyspnea score ^5^.

***Spirometric measurements***

At both visits, spirometry was performed before and after administration of 180 mcg of inhaled albuterol (ndd Easy-One spirometer, Andover, MA). Percent predicted values were calculated using Hankinson NHANES reference equations ^6^. COPD was defined by post-bronchodilator FEV_1_/FVC<0.70 at baseline visit per the GOLD guidelines ^3^. Bronchodilator responsiveness was defined as an increase in FEV_1_ or FVC by 200 mL and 12% from baseline. Disease severity was described by GOLD spirometric stage. “GOLD 0” was defined as post-bronchodilator FEV_1_/FVC≥0.70 at baseline visit and FEV_1_ percent predicted ≥80%. Participants with FEV_1_/FVC≥0.70 but with FEV_1_<80% predicted were considered to have Preserved Ratio Impaired Spirometry (PRISm) ^7^.

***CT measurements***

Using 3D Thirona software (www.thirona.eu), emphysema was quantified as the percentage of lung voxels with attenuation lower than -950 HU at maximal inspiration (%LAA-950) at Visit 1 and Visit 2 ^8^. The ratio of lung upper third to lower third emphysema %LAA-950 was used to evaluate the apico-basal emphysema distribution *(ratio950*). The Hounsfield units at the 15^th^ percentile of the CT density histogram at end-inspiration using Thirona software corrected for the variations in depth of inspiration *(Adjusted Perc15)* were used in the analyses of longitudinal changes in emphysema, as this may be a more robust measure of emphysema progression ^9,10^. Airway disease was assessed using VIDA software (www.vidadiagnostics.com) as gas trapping (percentage of low attenuation units less than -856HU at end-expiration), airway wall thickness (obtained along the center line of the lumen, in the middle third of the airway segment, for one segmental airway of each lung lobe; the mean value across all lobes was used for analysis), and Pi10 (the square root of the wall area of a hypothetical airway of 10-mm internal perimeter).

**Variance due to measurement error of ∆FEV_1_**

Consider the measured difference in FEV_1_ from Visit 1 to Visit 2. Assuming the measured outcome FEV_1_ is comprised of the true value of FEV_1_ and measurement error, the variance of ∆FEV_1_ can be written in terms of the true measurement and measurement error as follows:

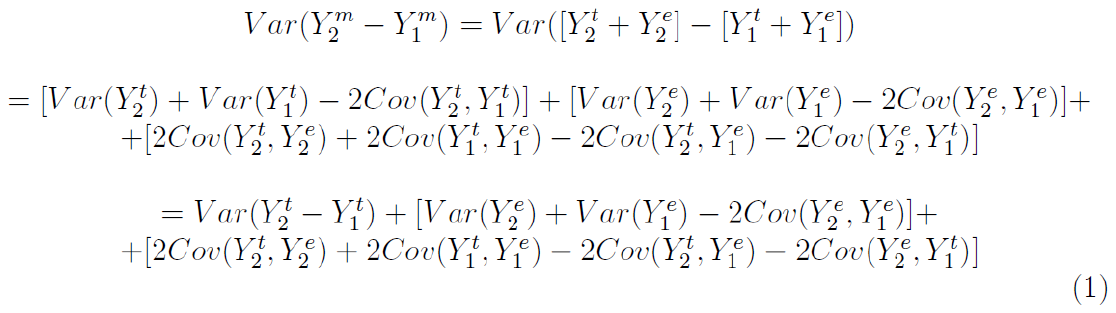

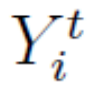

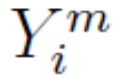

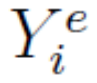
where denotes FEV_1_ measured at Visit *i*; denotes the true value of FEV_1_ at Visit *i*; and denotes the measurement error associated with the measured value of FEV_1_ at Visit *i* for *i* = 1 ; 2. Equation 1 assumes that the measured outcome FEV_1_ is comprised of the true value of FEV_1_ and measurement error such that *i* = 1 ; 2.

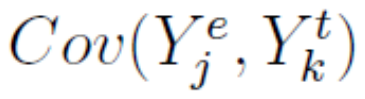
Assuming the true measurement is independent of the measurement error, then the covariance between the true measurement and measurement error is zero (i.e. = 0 for *j , k* = 1 ; 2). Then,

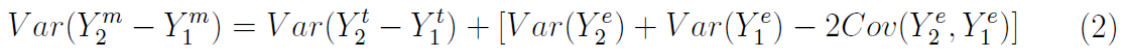

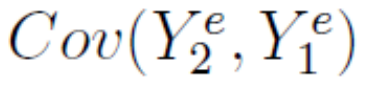
If we assume further that the measurement error associated with FEV_1_ at Visit 1 is independent of the measurement error associated with FEV_1_ at Visit 2, then = 0 and we can rewrite equation 2 as follows:

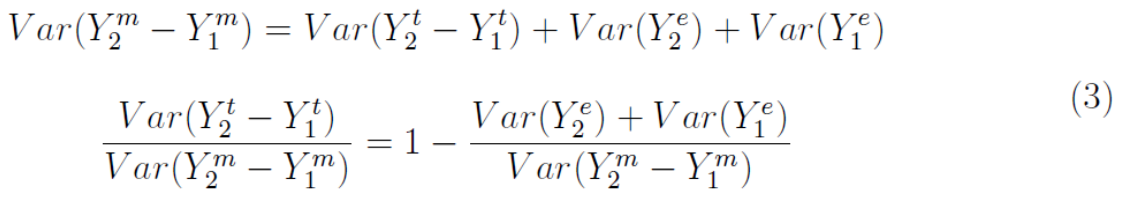

From existing literature ^11^, the coefficient of variation associated with repetitive measurements of FEV_1_ over a short period of time in patients with obstructive lung disease was shown to be ~0.04-0.3% over a wide range of FEV_1_. In the COPDGene study,

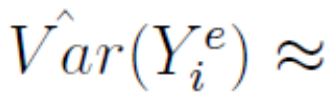
 10,000 for *i* = 1 ; 2

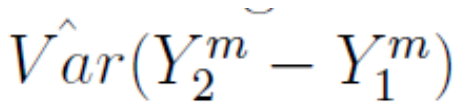
 = 90,000.

Then,

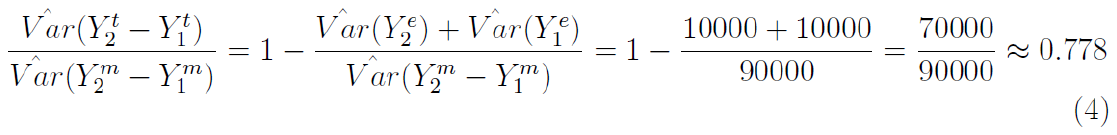

Based on the assumptions made above, we expect 22.2% of the variance of ∆FEV_1_ to be due to measurement error in FEV_1_.
